# Respiratory human adenovirus outbreak captured in wastewater surveillance

**DOI:** 10.1101/2024.06.15.24308982

**Authors:** Katherine R. Kazmer, Michelle L. Ammerman, Elizabeth A. Edwards, Marisa C. Eisenberg, Julie Gilbert, JoLynn P. Montgomery, Virginia M. Pierce, Jason B. Weinberg, Krista R. Wigginton

**Author notes:** Corresponding author Mailing address: Department of Civil and Environmental Engineering, 1351 Beal Ave., 181 EWRE, Ann Arbor, MI 48109-2125, USA.

## Abstract

Adenoviruses present challenges for traditional surveillance methods since there are more than 60 types that infect humans. Wastewater-based surveillance can supplement traditional surveillance methods for gastrointestinal-associated adenoviruses, but the ability to detect trends of respiratory-associated adenoviruses in wastewater remains unclear. We quantified human adenovirus type 4 (HAdV-4) in wastewater settled solids and compared wastewater measurements to clinical cases from an outbreak investigation beginning in late September 2023. The human adenovirus type 4 target was positively correlated with clinical cases (Spearman’s rho = 0.5470, p < 0.0001) and followed a similar trend during the outbreak. We also quantified human adenovirus types 3, 7, 14, 21, 40/41, and a pan-adenovirus assay that targets all types that infect humans. The respiratory adenoviruses comprised a small fraction of the adenoviruses in wastewater and types 40/41, which typically cause gastrointestinal disease, comprised the majority of the detected adenoviruses. The efficacy of adenovirus wastewater surveillance will depend on assay specificity and the public health action available for adenovirus types.

**WATER IMPACT STATEMENT:** Community adenovirus surveillance is strengthened by wastewater measurements. We evaluated the correlation between wastewater measurements and clinical cases during an outbreak of respiratory adenovirus type 4 infections on a college campus. Results indicate respiratory adenoviruses comprise a small portion of the adenoviruses measured in wastewater and the utility of adenovirus wastewater surveillance depends on the type and public health actionability.

**GRAPHICAL ABSTRACT:** 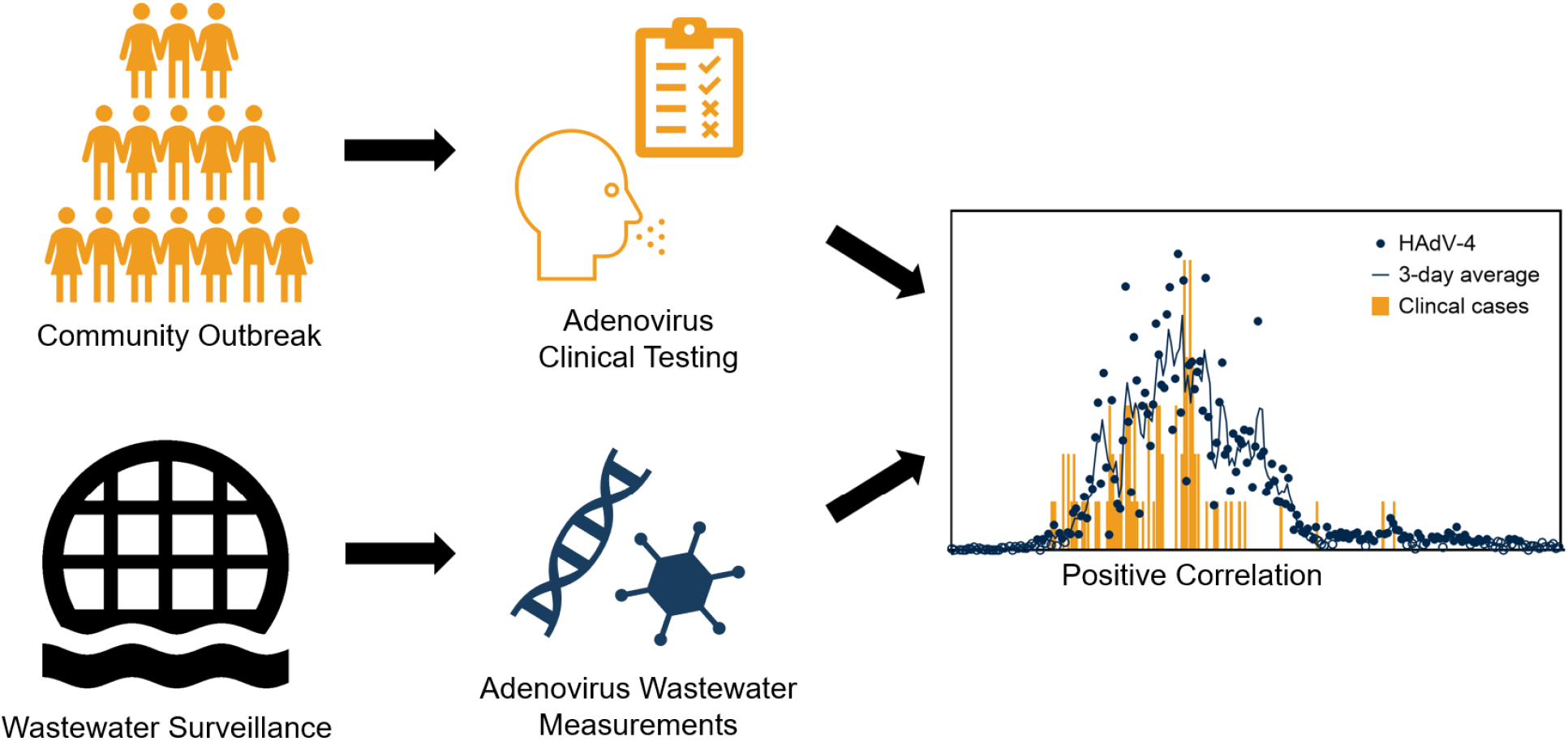

## INTRODUCTION

Adenoviruses are non-enveloped double-stranded DNA viruses of the family *Adenoviridae*. There are over 100 known adenovirus types and >60 that infect humans categorized in 7 species.^1^ Each species can cause a variety of illnesses including respiratory tract infections, gastroenteritis, ocular disease, and cystitis, although concurrent infections with multiple types of human adenovirus (HAdV) are rare.^2^ Adenovirus types 40/41 (species F) are commonly associated with gastrointestinal illness and species B, C, and E are commonly associated with respiratory illness. Adenovirus transmission can occur through the environment via respiratory droplets or fecal material, and adenovirus virions can remain infectious for long periods of time outside of their host.^3^ Adenoviruses are one of three viruses that are listed on the U.S. Environmental Protection Agency’s (EPA) Contaminant Candidate List (CCL 5) due to their persistence in water treatment. There are no approved antivirals or therapeutics to treat patients with adenovirus infections. An oral live virus vaccine covering types 4 and 7 has been developed but is not universally available.^4^

The frequency of asymptomatic cases combined with the lack of mandatory reporting, of clinical tests used as frequently as other respiratory viruses, and of labs that sequence clinical samples regularly for adenovirus typing, makes it difficult to detect adenovirus outbreaks and identify seasonal trends. It has been estimated that 40% of adenovirus cases are asymptomatic,^5^ and there is some evidence suggesting winter seasonality.^1,6–8^ Although it is not mandatory to report adenovirus cases in the U.S., passive surveillance from 2003-2016 indicated that type 3 (species B) was the most reported followed by types 2 and 1 (both species C).^1^

Wastewater surveillance provides an opportunity to shed light on the trends of HAdV infections within and between communities. Prior to COVID-19, most research on the detection and quantification of adenoviruses in wastewater focused on informing wastewater treatment and mitigating waterborne transmission. That work focused primarily on types 40/41 due to the fact that they are associated with gastrointestinal illness and are shed during and after an infection and detected in wastewater,^5^ although respiratory adenoviruses were also detected in wastewater residual solids.^7^ More recently, efforts have shifted to detecting and quantifying adenoviruses in wastewater for epidemiological purposes.^9–11^ The Wastewater Sewer Coronavirus Alert Network (WastewaterSCAN) network recently demonstrated a positive correlation between the concentrations of adenovirus types 40/41 in wastewater primary solids and clinical data.^12^ Adenovirus species B and C have also been detected in wastewater;^6,8,9^ however there is still a lack of evidence that respiratory HAdV levels in wastewater correlate with clinical cases. The signals of many other respiratory viruses have been measured in wastewater and correlated with clinical cases.^8,10,11,13–17^ Wastewater surveillance of respiratory-associated adenoviruses has the potential to help inform public health actions aimed at reducing transmission and inform the medical community of possible sources of respiratory illnesses.

In September 2023, Ann Arbor, MI experienced an outbreak of respiratory infections caused by adenovirus type 4.^18,19^ This outbreak provided an opportunity to study the relationship between wastewater levels and confirmed cases for a respiratory adenovirus. We retroactively measured adenovirus type 4 (species E) in stored samples collected from the City of Ann Arbor Wastewater Treatment Plant. In addition to type 4, we also measured adenovirus types 3, 7, 14, 21 (species B) and types 40/41 (species F), and we used a pan-adenovirus assay that targets all types that infect humans. Ultimately, this work demonstrates the feasibility of detecting respiratory adenovirus outbreaks in wastewater despite the relatively small signal that respiratory adenoviruses comprise compared to gastrointestinal adenoviruses.

## MATERIALS AND METHODS

### Sample Collection and Preparation

We focused on primary settled solids rather than wastewater influent as an early experiment suggested that a common method for detecting viruses in primary settled solids was more sensitive than a common method for detecting viruses in composite influent samples (Figure S1). We describe methods used to measure targets in the primary solids below. Methods used for influent samples are described in the Supporting Information.

Samples of primary settled solids were collected daily from primary clarifiers at the City of Ann Arbor Wastewater Treatment Plant in 50 mL conical tubes and stored at 4 °C. Samples were transported 2x weekly to the University of Michigan campus. The treatment plant services a population of approximately 120,000 with a maximum and average flow capacity of 29.5 million gallons per day (MGD) and 18.5 MGD, respectively. The estimated solids retention time (SRT) in the primary clarifiers based on data from the treatment plant was 8 days. Samples were pasteurized at 56°C for 30 minutes and aliquoted into two 2 mL cryotubes. The aliquots were preserved at −80°C until further analysis.

### Nucleic Acid Extraction

Aliquots from August 2022 to March 2023 were thawed and extracted in batches in January 2024. Six dates were missing sample aliquots including 8/25/22, 9/12/22, 1/24/23, 1/26/23, 2/12/23, and 3/27/23. Once thawed, samples were centrifuged at 4000xg for 15 minutes to concentrate the solids. The supernatant was discarded, and the remaining pellet was used for nucleic acid extraction. For the nucleic acid extraction, 55.6-58.6 mg of the pelleted solids were combined in sterile 2 mL screw cap tubes (Fisher, Cat. No. 02-682-558) with 0.5 g of 0.5 mm zirconia/silica beads (BioSpec, Cat. No. 11079105z) that were previously sterilized at 400°C for 4 hours and 1.44 mL of DNA/RNA Shield (Zymo, Cat. No. R1100-250). The sample tubes were then vortexed briefly and chilled overnight at 4°C.

Following refrigeration, samples were homogenized in a BioSpec Bead Beater 96 for 2 minutes at 1000 rpm and centrifuged briefly at 17,000xg for 6 seconds to collect the beads. The supernatant of each sample was extracted in triplicate with a Kingfisher MagMAX™ Viral/Pathogen II Nucleic Acid Isolation Kit on a Kingfisher Flex System (Fisher, Waltham, MA). Extractions followed the MagMAX™ protocol. In brief, sample plates were prepared with 275 µL binding beads, 200 µL samples, and 5 µL proteinase K added to each well. Wash plates were prepared with 500 µL wash solution and 500 µL of 80% ethanol in each well. Sample extracts were eluted into 50 µL elution buffer and immediately analyzed by droplet digital PCR and RT-PCR. We tested inhibition by diluting sample extracts 1:40 and comparing gene copies from undiluted extracts to estimated gene copies from diluted extracts accounting for the dilution. The results suggested that fewer than 5% of undiluted samples had inhibition that resulted in measurements off by >2x (Figure S2). Sample extracts were diluted 1:40 for the targets that were in high abundance (i.e. HAdV-pan, HAdV-40/41, crAssphage, PMMoV). Sample extracts were not diluted for the HAdV-3, 4, 7, 14, and 21 targets to maximize assay sensitivity.

### Solids Characterization

The remaining solids from the pelleted sample (70.1-708.3 mg) were used for dry weight analysis. Solids were weighed and baked in aluminum trays at 105°C overnight in accordance with methods published previously.^10,16^ After drying, solids were weighed and the dry weight was calculated. Gene targets were reported as gene copies per dry weight of sample.

### Droplet Digital PCR and RT-PCR

Nucleic acid extracts were analyzed immediately for adenoviruses, pepper mild mottle virus (PMMoV), and crAssphage (recently renamed *Carjivirus*). A single wastewater sample resulted in three sample extracts and three wells of a PCR plate. Results from triplicate sample extracts were merged for data analysis using the sum of the positive droplets across triplicate wells. Each plate included three negative extraction controls that consisted of nuclease-free water that was extracted alongside samples and three non-template controls that consisted of nuclease-free water added to the PCR reaction in place of the template. Plates were rerun if either of the negative controls amplified above the threshold (merged positive droplets > 6). Positive controls were included on each plate and consisted of gBlocks obtained from Integrated DNA Technologies (IDT, Coralville, IA) for HAdV types 4, 14, 21, 40, 41, and pan-adenovirus, and Zeptometrix NATtrol™ Adenovirus Type 3 Stock, and Zeptometrix Adenovirus Type 7A Lysate (Zeptometrix, Buffalo, NY). The gBlock sequences, gene locations, and GenBank accession numbers are provided in Supporting Information Table S1. On each plate, all positive controls were amplified by the HAdV-pan assay and each type-specific adenovirus assay only amplified the matching type-specific positive control. Assays used to target specific viruses and adenovirus types are detailed in Table 1. Samples were re-extracted if the PMMoV target was not amplified and plates were rerun if positive controls did not amplify. Each PCR reaction (22 µL) consisted of 5.5 µL extracted nucleic acid template, 900 nM primers, 250 nM probe, 2.2 µL Reverse Transcriptase, 1.1 µL DTT, 5.5 µL Supermix, and remaining volume of nuclease-free water. Reverse Transcriptase, DTT, and Supermix were purchased together in the One-Step RT-ddPCR Advanced Kit for Probes (Bio-Rad Cat. No. 1864022). A control experiment was conducted in which 11 samples were analyzed using ddPCR Supermix for Probes (no dUTP) for DNA targets. This resulted in similar gene copies for adenovirus targets HAdV-3, HAdV-4, and HAdV-40/41 as when analyzed with RNA Supermix (Figure S3). A control experiment was conducted to characterize the degradation of targets during storage and freeze/thaw cycles. This result demonstrated that similar gene copies were observed for adenovirus targets HAdV-3 and HAdV-40/41 after two freeze/thaw cycles that occurred during storage for 2 months (Figure S4).

**Table 1.**
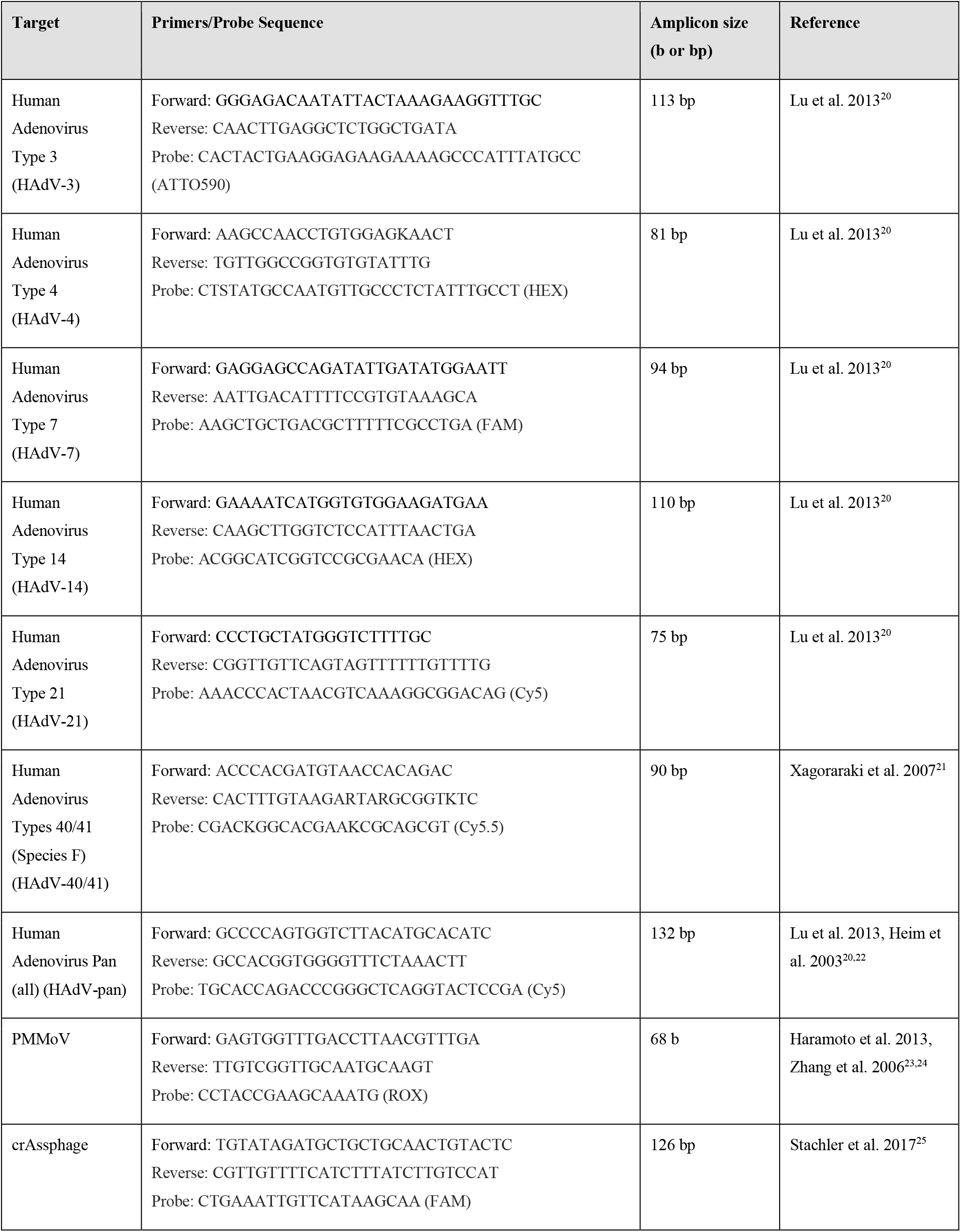
ddPCR targets and assay information.

Droplets were generated with a Bio-Rad Droplet Generator along with Droplet Generator Oil and Droplet Generator Cartridges. After droplets were generated, the 20 µL reactions were run on a thermal cycler with the following conditions: initial RT step at 50 °C for 60 min., initial denaturation at 95 °C for 5 min., 40 cycles of denaturation at 95 °C for 30 sec., annealing at 56 °C for 1.5 min., enzyme deactivation at 98 °C for 10 min., and a final hold at 4 °C. Within 24 hours of amplification, droplets were read on a Bio-Rad QX600™ Droplet Reader. Thresholds were set manually based on positive controls and separation between positive and negative droplets. Reactions with fewer than 10,000 droplets were removed from the data set and all other triplicate wells were merged for further data analysis. All negative extraction controls and non-template controls had fewer than 2 positive droplets. Merged wells with 6 or fewer positive droplets were considered below the assay’s limit of detection.

Target gene copies per gram of dry weight were calculated by dimensional analysis using the elution volume in the extraction and the measured dry mass of the solids. Gene copies were normalized by dividing the adenovirus target gene copies by PMMoV or crAssphage gene copies.

### Clinical Data

HAdV-4 cases were identified as part of an outbreak investigation by the University of Michigan University Health Service (UHS) and sequenced by the United States Centers for Disease Control and Prevention (CDC). Details of the outbreak investigation as well as the outbreak case definition have been published previously.^18,19^ Briefly, public health officials surveilled results from multi-pathogen respiratory panel testing for university students between the ages of 17 and 26 who presented to UHS or the university-affiliated emergency department (ED) with flu-like symptoms between 9/28/2022 and 1/30/2023. Cases were included if samples tested positive for adenovirus with the BioFire® Respiratory 2.1 (RP2.1) Panel (RPAN, BioFire Diagnostics, Salt Lake City, UT). This multiplex PCR panel targets 19 respiratory pathogens, including human adenoviruses but does not distinguish individual types. A total of 90 adenovirus cases were confirmed with the panel. A subset of 36 specimens were submitted to the CDC for typing and were determined to be HAdV-4 with 100% sequence homology in hypervariable regions 1-6. Clinical cases are reported based on specimen collection date from infected persons. Since only students aged 17 to 26 years were included in the outbreak case definition, for the analysis in this paper incidence rates were determined based on a student population of 52,000 during the study period.

In addition to the HAdV-4 cases in University of Michigan students described above, we obtained results from all clinical respiratory pathogen PCR panel (RPAN) and gastrointestinal pathogen PCR panel (GIPAN) tests conducted by the Michigan Medicine Clinical Microbiology Laboratory during the study period, including tests from cases in the outbreak. These results represent patients primarily located in the Ann Arbor area who presented at the hospital, but also include patients who presented at approximately 14 clinics in the Southeast Michigan region. The BioFire® FilmArray® Gastrointestinal (GI) Panel (GIPAN, BioFire Diagnostics, Salt Lake City, UT) targets 22 gastrointestinal pathogens including adenoviruses (types 40/41). All identifiers were removed, and the data were cleaned by removing duplicates and unknown or “test” values. Use of the human adenovirus case data was reviewed and approved by the University of Michigan Institutional Review Board (HUM00239913) and comparisons between UHS data and wastewater data were exempt.

### Statistical Analysis

All statistical analyses were performed using GraphPad Prism (La Jolla, CA) Version 10.1.2. with the exception of Kendall’s tau correlation which was performed in R Version 4.0.2 and RStudio Version 2023.12.1 Build 402.

## RESULTS AND DISCUSSION

Following notification of the confirmed adenovirus type 4 outbreak, we analyzed 215 wastewater settled solid samples collected from August 2022 to March 2023 for HAdV-4. Human adenovirus type 4 was detected in 68.4% (147/215) of the samples in the study period, and concentrations aligned with the outbreak trend as determined by case data (Figure 1A). Specifically, in the wastewater data, the target was first detected in the sample collected on 9/22/2022 and then levels rose for several weeks. The concentration peaked on November 13th at 4.8×10^5^ gc/g-dw. The concentrations then decreased to low concentrations and stabilized in early January 2023. By comparison, the first confirmed HAdV-4 case in the campus outbreak occurred on 9/28/2022. RPAN tests were generally reserved for those with prolonged symptoms; it was therefore likely that this individual had contracted the illness days to weeks before this date. The numbers of cases increased until peaking at 6 cases per day on November 15th and 17th and then decreased. The last HAdV-4 case in the outbreak was reported on 1/30/2023. The fact that wastewater detections for the HAdV-4 target continued to occur until nearly 6 weeks after the last confirmed case in the outbreak may be in part due to prolonged shedding, which can take place for weeks to months for some adenoviruses.^1,18,26^ It may also be due to the fact that the outbreak study focused on the university community, whereas the corresponding wastewater treatment plant encompasses a much larger population. Our ddPCR assay was designed for HAdV-4, and all clinical samples from the outbreak matched 100% with HAdV-4 sequences.^18,19^ Ultimately, these results suggest that respiratory adenovirus outbreaks can be detected through wastewater surveillance with assays that are specific for the adenovirus type.

**Figure 1.**
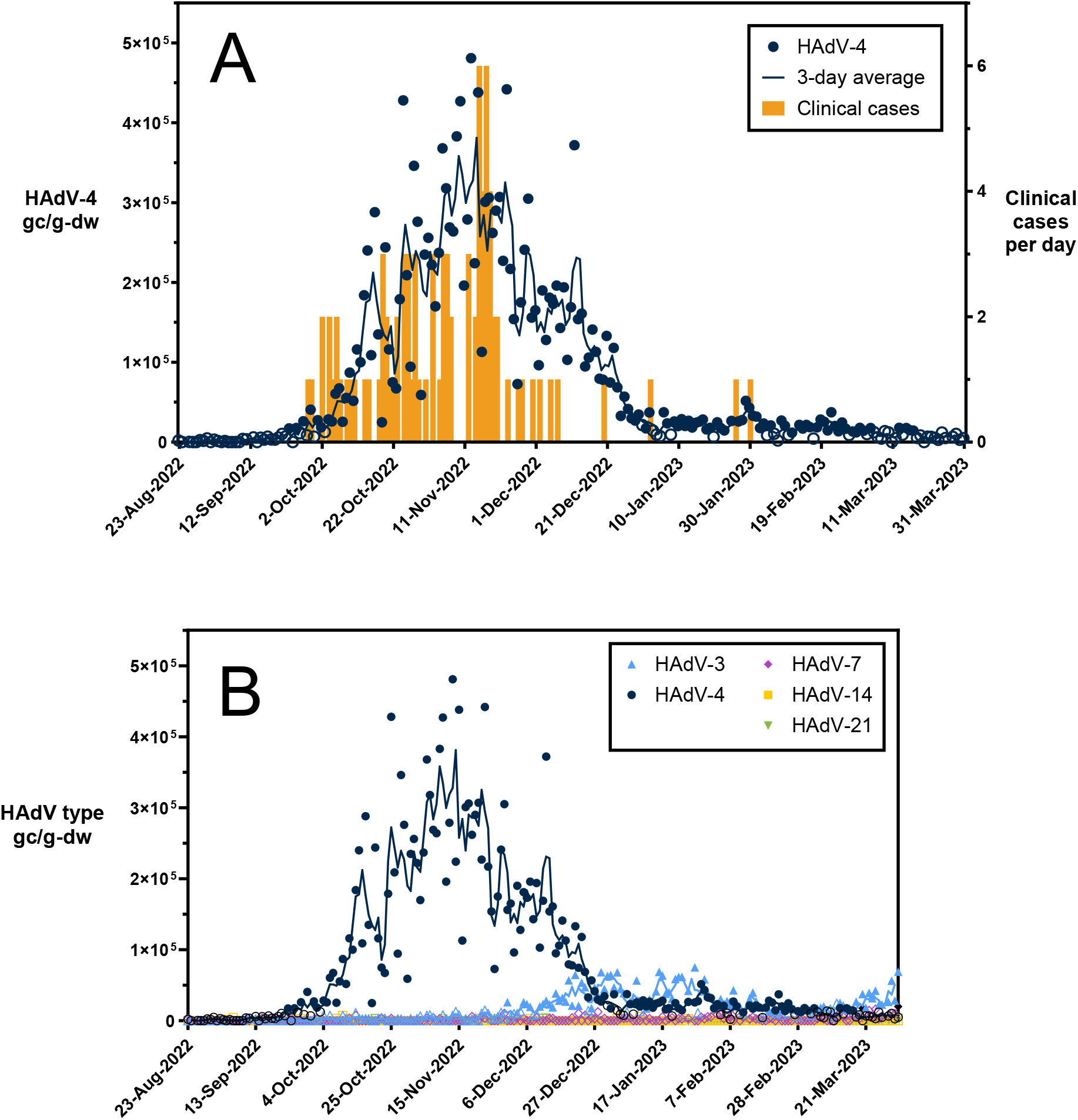
Respiratory human adenoviruses in wastewater during a HAdV-4 outbreak. Panel A shows HAdV-4 in wastewater solids and clinical cases per day. Panel B shows respiratory HAdV-3, 4, 7, 14, 21 in wastewater solids. HAdV concentrations in primary solids are reported as gene copies per gram dry weight. Open circles represent measurements that are below the assay limit of detection. Lines represent a three-day middle-centered average of the HAdV values. Clinical cases are reported as new cases per day that met the outbreak case definition as defined in Montgomery et al.^18^ Dates represent specimen collection dates for both wastewater primary solids and clinical specimens.

The trends in the wastewater data and the clinical cases were significantly correlated regardless of whether the primary solids concentrations were normalized by PMMoV or crAssphage concentrations. The wastewater data normalized with PMMoV had a slightly stronger correlation with clinical cases (Spearman’s rho = 0.5470, p < 0.0001) than non-normalized data (Spearman’s rho = 0.4507, p = 0.0015) and crAssphage-normalized data (Spearman’s rho = 0.3898, p = 0.0068) (Figure S5). Kendall’s tau analysis resulted in similar findings (τ = 0.301, p = 0.0077) and correlations are provided in Supporting Information Table S2.

Monitoring viruses in wastewater has been characterized elsewhere as a leading indicator compared to clinical data.^27–29^ In this study, HAdV-4 was first detected in the wastewater primary settled solids six days before the first case identified in the outbreak investigation on September 28th. However, the relatively small number of cases in this outbreak limits the ability to rigorously analyze leading or lagging signals. Additionally, it is worth noting that wastewater can only be a leading indicator in practice if the pathogen in question is regularly monitored, underscoring the importance of broadening the range of pathogens monitored in state and national programs if we are to take advantage of any practical lead time. In Influenza A outbreaks observed at the University of Michigan and Stanford, wastewater detection occurred at approximately the same time as the start of the outbreak based on confirmed cases.^15^ We note that the clinical course of HAdV is much longer than that of Influenza A.^30,31^ Data on fecal shedding levels and trajectories are limited for both IAV and respiratory HAdV at this time. The fact that HAdV-4 was not detected in samples before the outbreak suggests that it is not prevalent in wastewater outside of outbreaks. More studies will be necessary across multiple sewersheds and seasons to confirm that HAdV-4 is only occasionally detectable in wastewater.

To provide insight on the wastewater levels of respiratory human adenoviruses that were not associated with an outbreak, we also analyzed the samples for respiratory HAdV-3, HAdV-7, HAdV-14, and HAdV-21 (Figure 1B). We selected these adenovirus types because they cause epidemic acute respiratory infections along with HAdV-4.^1^ HAdV-3 was detected in 43.7% (94/215) of the wastewater solids samples. HAdV-3 was first detected on November 30th and then increased until peaking on January 27th at 7.5×10^4^ gc/g-dw. This concentration is approximately one order of magnitude lower than the peak HAdV-4 concentrations. HAdV-7 was detected in just three samples (3/215) at low levels on Feb. 2, Mar. 20, and Mar. 31. HAdV-14 and HAdV-21 were not detected in wastewater during the study period (0/215).

We compared the respiratory adenovirus levels detected in wastewater to clinical data generated from a respiratory pathogen PCR panel of 19 targets, including adenoviruses (RPAN). These types of respiratory pathogen PCR panels are used at many hospitals across the U.S. but are not quantitative and do not distinguish between human adenovirus types. While the RPAN incidence rate based on adenovirus positives is correlated with respiratory adenoviruses detected in wastewater (Spearman’s rho = 0.5703, p < 0.0001) during the study period, the RPAN adenovirus data and summed respiratory HAdV-3 and HAdV-4 wastewater concentrations did not align (Figure S6). Higher adenovirus incidence rates based on the RPAN results were observed during the HAdV-4 outbreak in November and December, but were also observed in late February and March when respiratory adenoviruses were not detected frequently in wastewater. The fact that the respiratory adenovirus trends did not align with RPAN incidence rate could be in part due to the fact that the sewershed population does not encompass the total RPAN testing population which includes patients who presented in Ann Arbor and at approximately 14 clinics in the Southeast Michigan region. It could also be due in part to respiratory adenovirus types that were not measured in wastewater but were encompassed in the RPAN assay (specific HAdV types measured by RPAN is proprietary information). The outbreak investigation team noted that adenovirus cases are likely underrepresented since adenovirus is not a notifiable disease and infections present with symptoms similar to those caused by other types of respiratory viruses.^18,19^

Previous efforts to quantify adenoviruses in wastewater for wastewater-based epidemiology have focused on both pan-adenovirus assays as well as assays focused on types that are associated with gastrointestinal illnesses (types 40/41).^12^ To understand how the levels observed for HAdV-4 compare with other circulating adenoviruses, we analyzed the same samples with a pan-adenovirus assay and an assay that targets HAdV-40/41. We detected human adenovirus types 40/41 and the targets captured with the HAdV-pan assay in all 215 samples (Figure 2). Overall, the trends and quantities observed with the HAdV-40/41 assay were similar to the quantities observed with the HAdV-pan assay. Concentrations of both targets were lowest in the late summer (August-September) and began increasing in October. The levels remained high through December and January and then began to decline in February. The highest value measured with the assay targeting HAdV-40/41 was 1.9×10^7^ gc/g-dw on January 25^th^, whereas the pan assay resulted in a similar concentration of 1.5×10^7^ gc/g-dw on the same day. Comparatively, we measured 2.7×10^4^ gc/g-dw of the HAdV-4 target on January 25th and concentrations of HAdV-4 were consistently ~2-3 log_10_ lower than concentrations measured with the HAdV-40/41 and the pan-adenovirus assays. At the peak of the HAdV-4 outbreak on November 13th, the measured concentrations with the HAdV-40/41 and pan-adenovirus assays were 8.9×10^6^ gc/g-dw and 5.9×10^6^ gc/g-dw, respectively. Comparatively, measured HAdV-4 concentrations were 4.8×10^5^ gc/g-dw. These results indicate that throughout our study period, nearly all the adenovirus signal in the wastewater primary solids samples consisted of type 40/41 (species F). Consequently, the pan-adenovirus assay would not have captured an increase in levels due to the HAdV-4 respiratory adenovirus cases.

**Figure 2.**
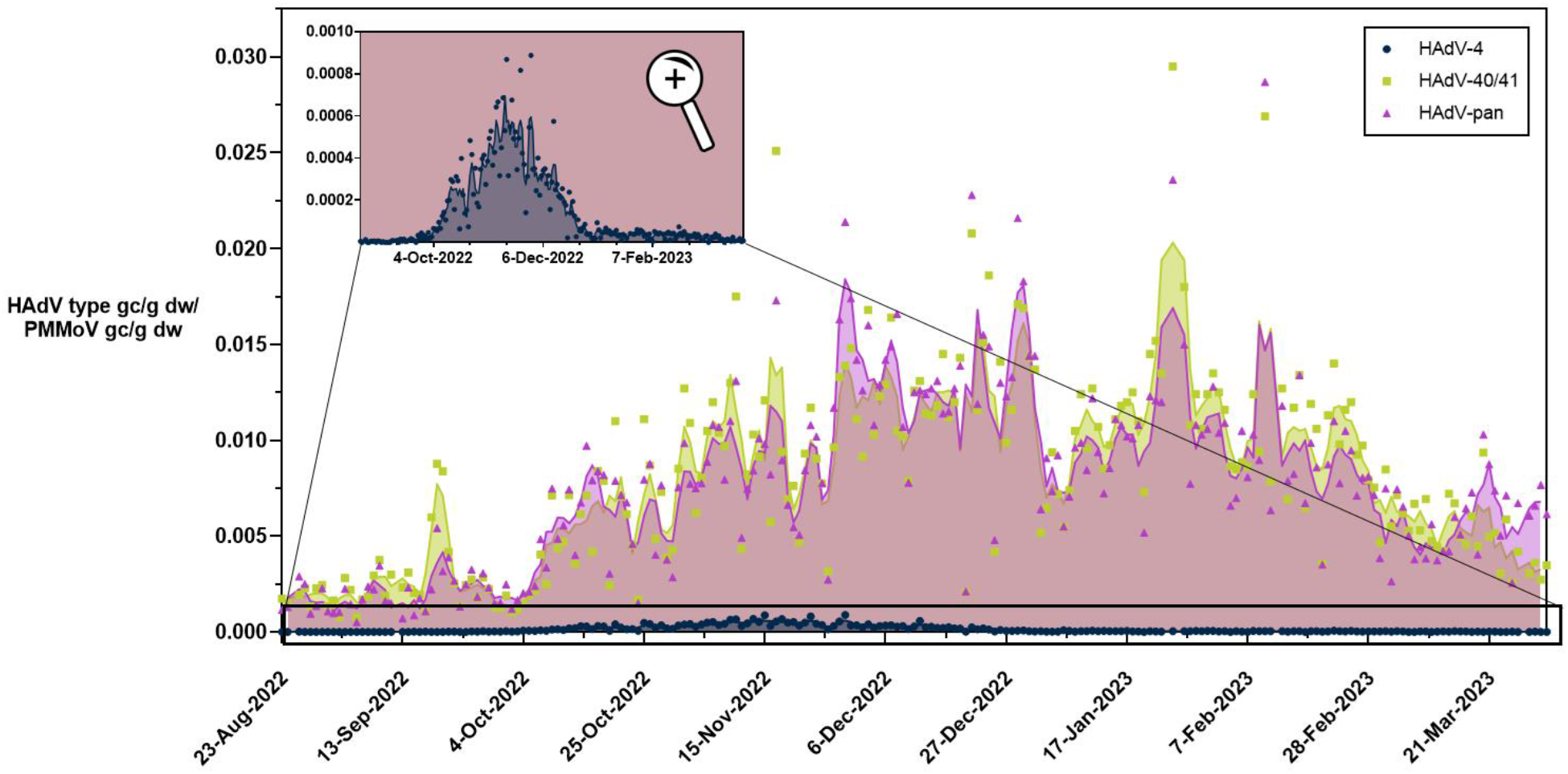
Human adenovirus targets HAdV-4, HAdV-40/41, and pan-adenovirus in wastewater primary settled solids. Concentrations are reported as target gene copies per PMMoV gene copies. Inset provides close-up of HAdV-4 outbreak over the same period. Open shapes represent measurements below the assay limit of detection. The lines represent a three-day middle-centered average of the normalized adenovirus target values. Dates represent wastewater specimen collection date.

Our HAdV-40/41 concentrations were in the range of levels reported in other studies. Boehm et al. reported median HAdV-40/41 concentrations of 5.8×10^5^ and 1.9×10^6^ gc/g-dw and maximum values of 5.8×10^6^ and 8.5×10^7^ gc/g-dw at two wastewater treatment plants in Southern California.^12^ Our results comparing levels of HAdV-4 in several paired settled solids and influent samples suggest partition coefficient (K_id_) values on the order of 1×10^4^ mL/g, similar to HAdV-40/41 K_id_ values reported previously.^12,32,33^ For reference, the reported K_id_ for SARS-CoV-2 is 1×10^3^ mL/g.^34^ Our results combined with past research suggest that wastewater solids may contain adenoviruses at concentrations 10,000 times those found in influent. As such, monitoring primary settled solids could be more effective at monitoring respiratory human adenoviruses due to their low concentrations.

We compared the gastrointestinal adenovirus levels detected in wastewater (HAdV-40/41) to clinical data generated from a gastrointestinal pathogen PCR panel of 22 targets (GIPAN) (Figure S6). The GIPAN included human adenovirus types 40/41. The GIPAN adenovirus incidence rate was not as highly correlated with gastrointestinal adenoviruses (Spearman’s rho = 0.2439, p = 0.0003). This is likely due to the low GIPAN test incidence rate during the 7-month study period. A previous study with a higher percentage of positive tests and longer study period demonstrated that HAdV-40/41 in wastewater correlated with GIPAN cases (τ = 0.30 and τ = 0.38, p < 10^−3^).^12^

Adenovirus outbreak investigations often rely on typing clinical specimens. The clinical data generated from respiratory panels are widely available but lack resolution to determine the specific adenovirus type causing an outbreak.^9,20^ Typing is typically achieved by sequencing and often only applied retrospectively when an outbreak is suspected. In this outbreak, typing helped confirm that all the respiratory adenovirus cases were related and caused by HAdV-4. Wastewater surveillance could provide a route to more efficiently detect and track respiratory adenovirus outbreaks. One approach would be to run a number of assays throughout respiratory illness season that target the different HAdV types. This, however, could be costly given the large number of HAdV types associated with respiratory illness. Wastewater sequencing approaches may also work for this purpose. Holland et al. recently applied sequencing to specify the relative abundances of different HAdV-4 and HAdV-7 in samples from manholes at a public university campus in Arizona.^9^ Our work suggests that quantitative PCR results from HAdV-pan assays would be unlikely to detect an outbreak of HAdV associated with respiratory illnesses given the high signals of HAdV associated with gastrointestinal illness. While management of patients with adenovirus infections does not vary based on adenovirus type, determination of adenovirus type is essential to identify an outbreak as early as possible. Early indication of adenovirus type through routine wastewater surveillance can detect and identify potential outbreaks earlier than clinical testing and justify allocation of resources towards outbreak prevention and reducing transmission.

There is continued interest in monitoring adenovirus in wastewater for public health purposes and adenovirus types 40/41 recently were included as a core target for the CDC National Wastewater Surveillance System (NWSS).^35^ Respiratory adenoviruses including HAdV-3 and HAdV-4 were not included on the list of core targets despite the fact that infections with respiratory adenovirus types are often more severe than those caused by gastrointestinal adenovirus types.^1,18–20,36^ Kujawski et al. reported outbreaks at 5 college campuses during the 2018-2019 academic year that were caused by respiratory adenoviruses and which led to 168 identified cases, 11 hospitalizations, and 2 deaths.^36^ The university outbreak we studied here included 7 otherwise healthy students with severe or critical illness caused by HAdV-4 infection, including 2 who required care in an intensive care unit.^18,19^

## CONCLUSION

There is a growing public health need for better surveillance of respiratory viruses in the environment since they can spread rapidly and manifest in severe illness. Respiratory adenoviruses have not been well-studied in wastewater. We demonstrate that wastewater measurements of respiratory HAdV can detect local outbreaks. Specifically, we show that clinical cases of infections caused by HAdV-4 correlated with HAdV-4 concentrations in primary settled solids from the local wastewater treatment plant. Our findings suggest that type-specific monitoring will be necessary to detect respiratory adenovirus outbreaks, as the pan-adenovirus assay results were dominated by HAdV-40/41 (species F), which do not typically cause outbreaks of respiratory illness.

We note that our results were limited to a single sewershed and a 7-month study period. Additional studies should be conducted in different sewersheds and across seasons to further demonstrate correlations between respiratory adenoviruses and clinical cases. The trends we observed in the HAdV-4 outbreak in the local community may be different than trends observed in other communities with different population sizes, sewershed dynamics, and individual shedding rates. Studies on fecal and urine shedding quantities and trajectories of various HAdV types would help in the interpretation of HAdV wastewater trends and applications of the wastewater data. Our research demonstrates that HAdV-40/41 dominates the wastewater environment and that specific targeted efforts to develop and monitor a panel of respiratory adenovirus or individual types are needed to optimize detection of local outbreaks in the community. Ultimately, these results will pave the way for future adenovirus surveillance and advance wastewater-based epidemiology and public health efforts to curb outbreaks.

## Data Availability

All data produced in the present study are available upon reasonable request to the authors.

