## Supplemental Information for "Respiratory human adenovirus outbreak captured in wastewater surveillance"

*Corresponding author

Number of Pages: 11

Number of Tables: 2

Number of Figures: 6

**Table of Contents**

*Figure S1.* Comparison of HAdV-4 in Influent and Primary Solids…………………………...3
*Figure S2.* ddPCR Inhibition Control………………………………………………………..4
Influent Sample Collection and Processing…………………………………………………….5
*Table S1.* Sequence of gBlock standards, gene location, and GenBank accession numbers for adenovirus targets…………………………………………………………………………….....6
*Figure S3.* Comparison of RNA and DNA Supermix………………………………………….7
*Figure S4.* Comparison of HAdV-3 and HAdV-40/41 gene copies before and after two freeze/thaw cycles……………………………………………………………………………………….....8
*Figure S5.* Correlation between clinical cases and non-normalized HAdV-4, HAdV-4 normalized by PMMoV, and HAdV-4 normalized by crAssphage.………….……………………………….9
*Table S2.* Kendall's tau correlation values…………………………………………………....10
*Figure S6.* Comparison between wastewater and RPAN/GIPAN data…………………………11

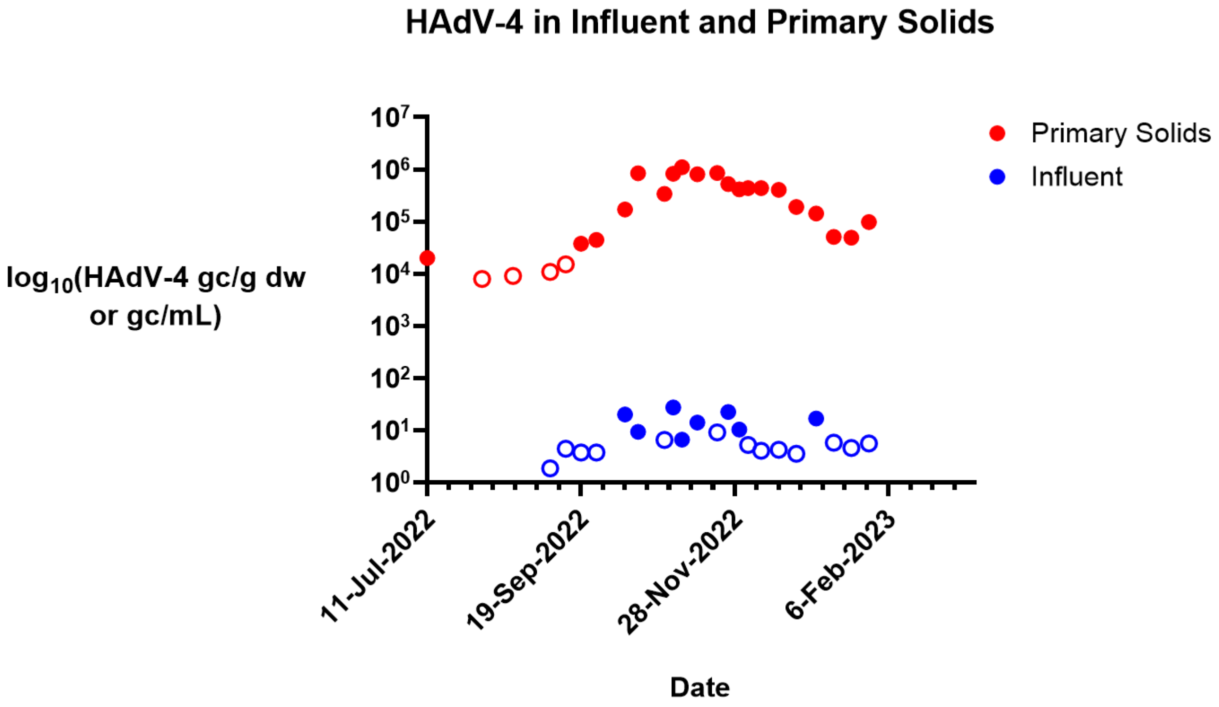


*Figure S1*. Comparison of HAdV-4 in Influent and Primary Solids. Influent samples shown in blue circles and primary solids samples shown in red circles. Open circles indicate the targets were below the limit of detection (LOD) and are plotted at the value below the LOD for that assay. The y-axis values are reported as gene copies per gram dry weight or gene copies per milliliter and are $\log_{10}$-scaled for convenience.

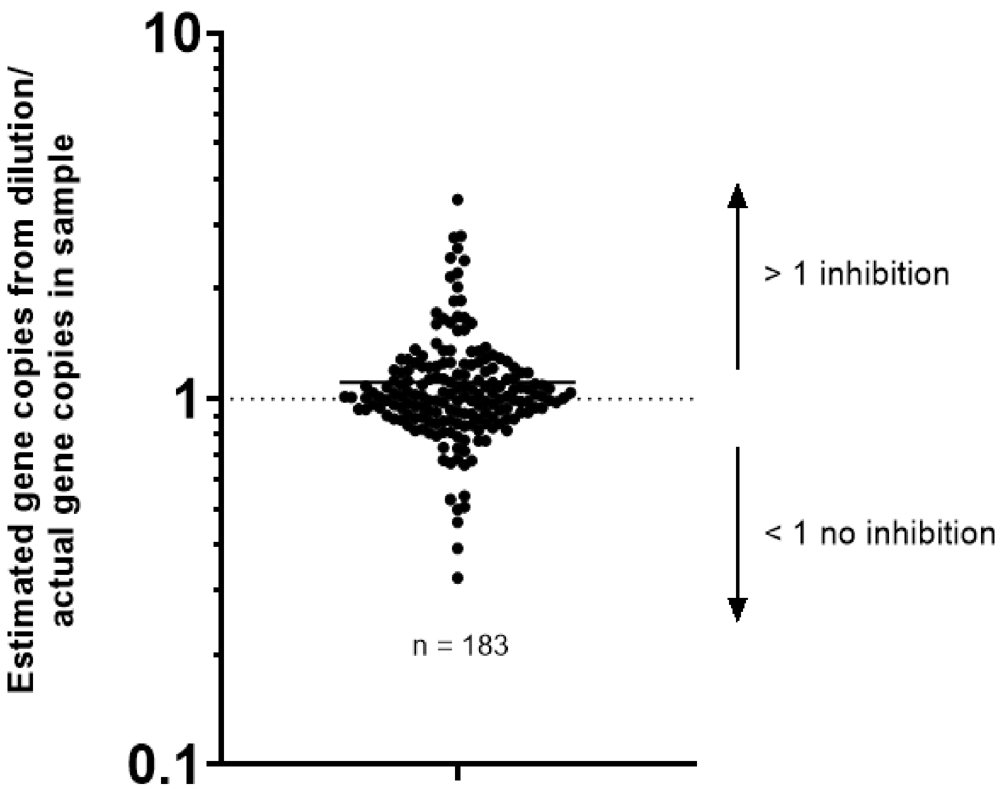


*Figure S2.* ddPCR Inhibition Control. 183 primary solids samples were diluted 1:40 and measured for adenovirus target HAdV-40/41. Diluted samples were compared to undiluted samples and the ratio of the estimated gene copies from the dilution divided by the actual gene copies in the undiluted sample is plotted. Ratios >1 represent inhibition from the sample disrupting the assay while ratios <1 represent little to no inhibition from the samples. The geometric mean for all values is plotted as a horizontal black bar at 1.108.

**Influent Sample Collection and Processing**

24-hour time-weighted composite influent samples were collected daily at a treatment plant in Southeast Michigan in 50 mL conical tubes and stored at 4ºC and transported 2x weekly to the University of Michigan campus. Samples were pasteurized at 56ºC for 30 minutes. NaCl and PEG were added to reach final concentrations of 0.2 M NaCl and 8% PEG and mixed by inversion until all PEG was dissolved. Following PEG addition, samples were refrigerated at 4ºC for 4+ hours or overnight. Following refrigeration, the precipitated samples were centrifuged at 4,700xg for 45 minutes at 4ºC. A majority of the supernatant was removed and discarded, while the remaining pellet was resuspended with 1-5 mL supernatant kept in the bottom of each sample. The final sample volume was recorded and gene targets are reported as gene copies per mL. Concentrated influent samples were extracted similarly to settled solids samples as described in nucleic acid extraction methods and gene targets were amplified with ddPCR and RT-ddPCR.

*Table S1.* Sequence of gBlock standards, gene location, and GenBank accession numbers for adenovirus targets

| **Assay** | **gBlock sequence** | **Gene location** | **GenBank accession no.** |
|---|---|---|---|
| HAdV-4 | 5’ - TCC TTT TGC CAT GGA AAT CAA CAT CCA AGC CAA CCT GTG GAG GAA CTT CCT CTA TGC CAA TGT TGC CCT CTA TTT GCC TGA TAA ATA CAA ATA CAC ACC GGC CAA CAT CAC CCT GCC CAC CAA CAC C - 3’ | 19,567-19,693 | AY594253 |
| HAdV-14 | 5’ - GAT GTG CGT GTT ATT GAA AAT CAT GGT GTG GAA GAT GAA CTT CCC AAC TAC TGT TTT CCA CTG GAC GGC ATC GGT CCG CGA ACA GAT AGT TAC AAG GAG ATT CAG TTA AAT GGA GAC CAA GCT TGG AAA GAT GTA AAT CCA AAT GG - 3’ | 19,428-19,573 | AY803294 |
| HAdV-21 | 5’ - GTA TGG TGG TAG AGC TCT AAA GCC AGA AAC TAA AAT GAA ACC CTG CTA TGG GTC TTT TGC TAA ACC CAC TAA CGT CAA AGG CGG ACA GGC AAA ACA AAA AAC TAC TGA ACA ACC GCA AAA CCA GCA GGT TGA ATA TGA TAT TGA CAT G - 3’ | 19,107-19,254 | AY601633 |
| HAdV-40 | 5’ - CAA GTT CAG AAA CCC CAC CGT GGC TCC CAC CCA CGA TGT AAC CAC AGA CAG GTC GCA GCG ACT GAC GCT GCG CTT CGT GCC CGT CGA CCG CGA GGA AAC CGC CTA CTC TTA CAA AGT GCG CTT TAC GCT GGC CGT GGG CGA CAA CCG G - 3’ | 17,771-17,918 | NC_001454 |
| HAdV-41 | 5’ - CAA GTT CAG AAA TCC CAC TGT GGC TCC GAC CCA CGA TGT AAC CAC AGA CAG GTC ACA GCG ACT GAC GCT GCG ATT CGT GCC AGT CGA CCG CGA GGA CAC CGC TTA TTC TTA CAA AGT GCG CTT TAC GCT GGC CGT GGG CGA CAA CCG G - 3’ | 17,771-17,918 | NC_001454 |
| HAdV-pan | 5’ - AGA TGG CCA CCC CAT CGA TGA TGC CCC AAT GGG CAT ACA TGC ACA TCG CCG GAC AGG ATG CTT CGG AGT ACC TCA GTC CGG GTC TGG TGC AGT TCG CCC GTG CAA CAG ACA CCT ACT TCA GTA TGG GGA ACA AAT TTA GAA ACC CCA CAG TGG CGC CCA CCC ACG ATG TGA CCA - 3’ | 18,416-18,589 | NC_011203 |

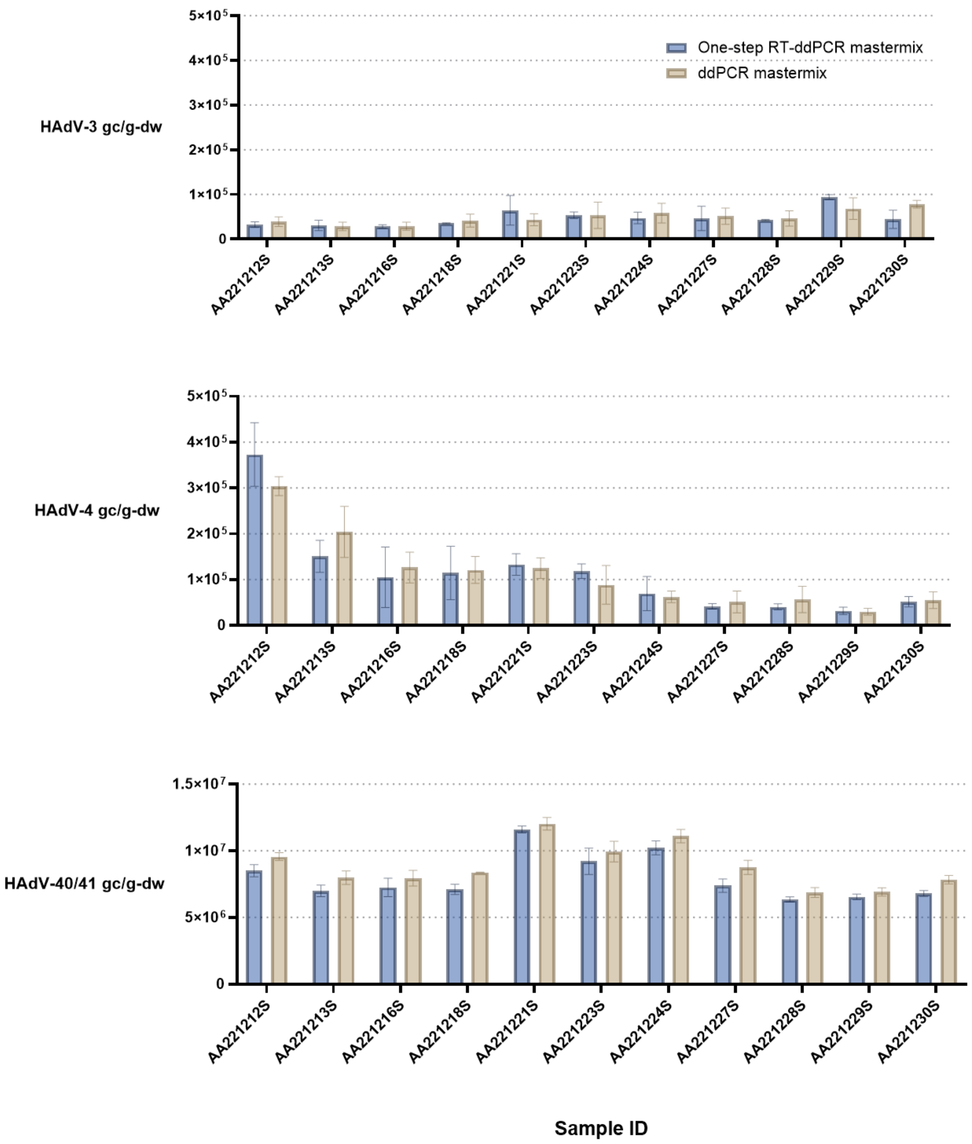


*Figure S3*. Comparison of RNA and DNA Supermix. Eleven samples are listed on the x-axis and were analyzed for three adenovirus targets: HAdV-3 (top), HAdV-4 (middle), HAdV-40/41 (bottom). Gene copies per gram dry weight are shown as bars in blue using one-step RT-ddPCR mastermix for RNA targets and in tan using ddPCR mastermix for DNA targets. Error bars represent the standard error of three independent sample replicates extracted and analyzed on the same day. Thermal cycler conditions varied since ddPCR mastermix for DNA targets did not need an initial reverse transcription (RT) step. All other thermal cycler conditions remained unchanged.

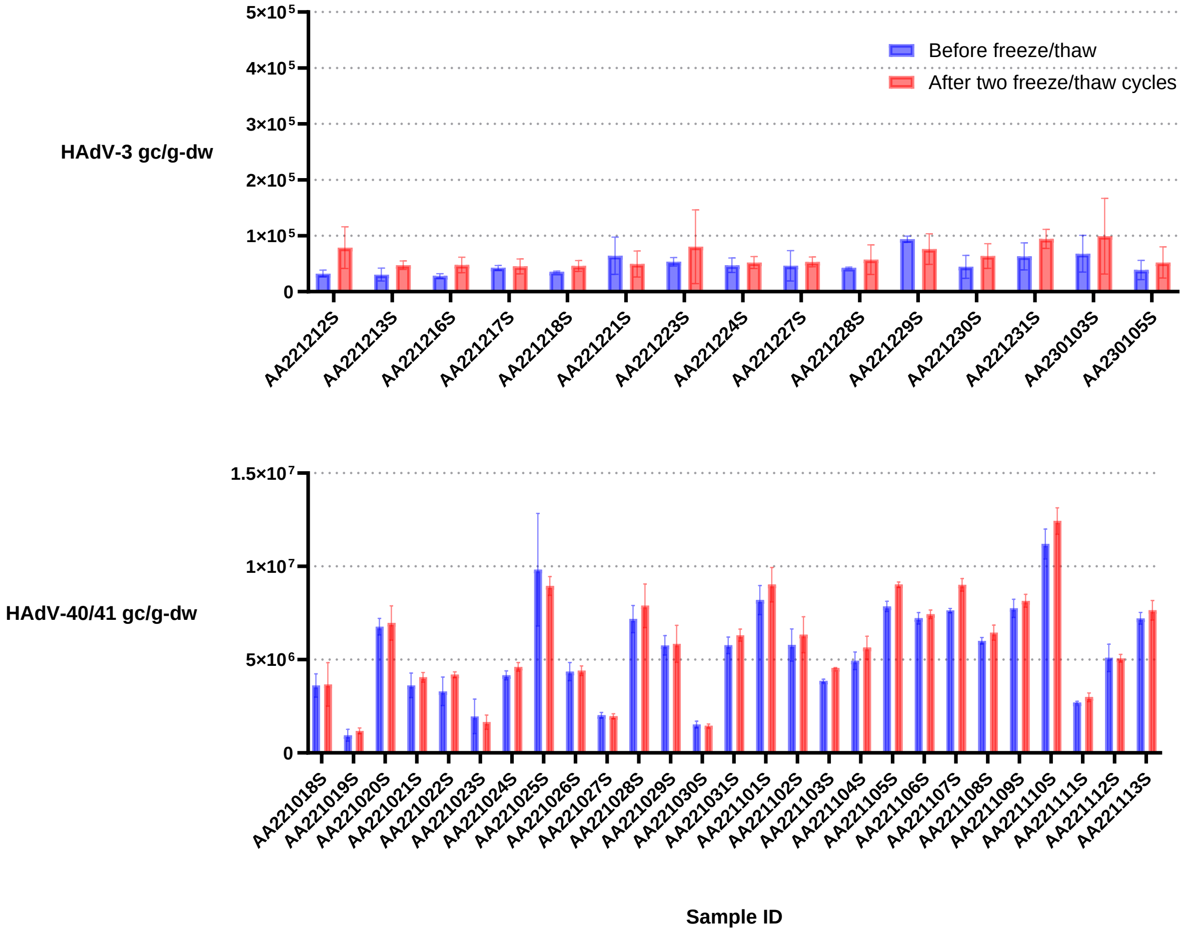


*Figure S4.* Comparison of HAdV-3 and HAdV-40/41 gene copies before and after two freeze/thaw cycles. Sample IDs are listed on the x-axis and were analyzed for two adenovirus targets: HAdV-3 (top) and HAdV-40/41 (bottom). Gene copies per gram dry weight are shown as bars in blue quantified before freeze/thaw and in purple quantified after two freeze/thaw cycles. Error bars represent the standard error of three independent sample replicates extracted and analyzed on the same day.

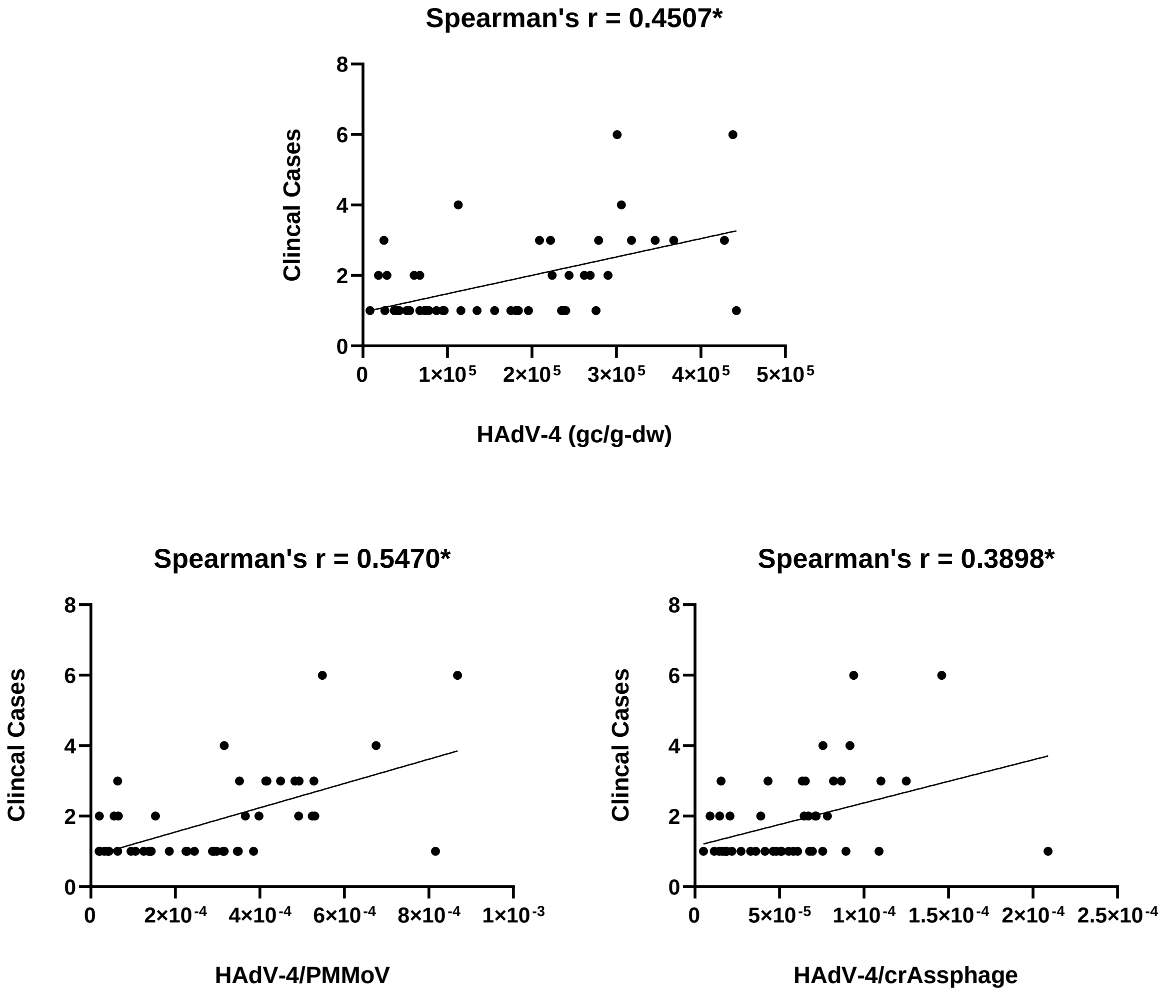


*Figure S5.* Correlation between clinical cases and non-normalized HAdV-4, HAdV-4 normalized by PMMoV, and HAdV-4 normalized by crAssphage. The clinical cases as described in the outbreak in Figure 1 were correlated with HAdV-4 in gene copies per gram dry weight (top), HAdV-4 normalized with PMMoV gene copies (bottom, left), and HAdV-4 normalized with crAssphage gene copies (bottom, right). The Spearman’s correlation coefficient was calculated and is noted on the plot. All correlation coefficients were significant.

*Table S2.* Kendall's tau correlation values

| | HAdV-4 | HAdV-4/PMMoV | HAdV-4/crAssphage |
|---|---|---|---|
| Clinical cases in outbreak | **τ = 0.301, p = 0.0077** | **τ = 0.425, p = 0.0001** | **τ = 0.309, p = 0.0061** |

| | 3-day middle-centered average HAdV-3 + 4 | 3-day middle-centered average HAdV-3 + 4/ PMMoV | 3-day middle-centered average HAdV-3 + 4/ crAssphage |
|---|---|---|---|
| 3-day middle-centered average RPAN incidence rate | **τ = 0.414, p < 0.0001** | **τ = 0.422, p < 0.0001** | **τ = 0.408, p < 0.0001** |

| | 3-day middle-centered average HAdV-40/41 | 3-day middle-centered average HAdV-40/41/ PMMoV | 3-day middle-centered average HAdV-40/41/ crAssphage |
|---|---|---|---|
| 3-day middle-centered average GIPAN incidence rate | **τ = 0.185, p = 0.0005** | **τ = 0.195, p = 0.0002** | **τ = 0.206, p = 0.0001** |

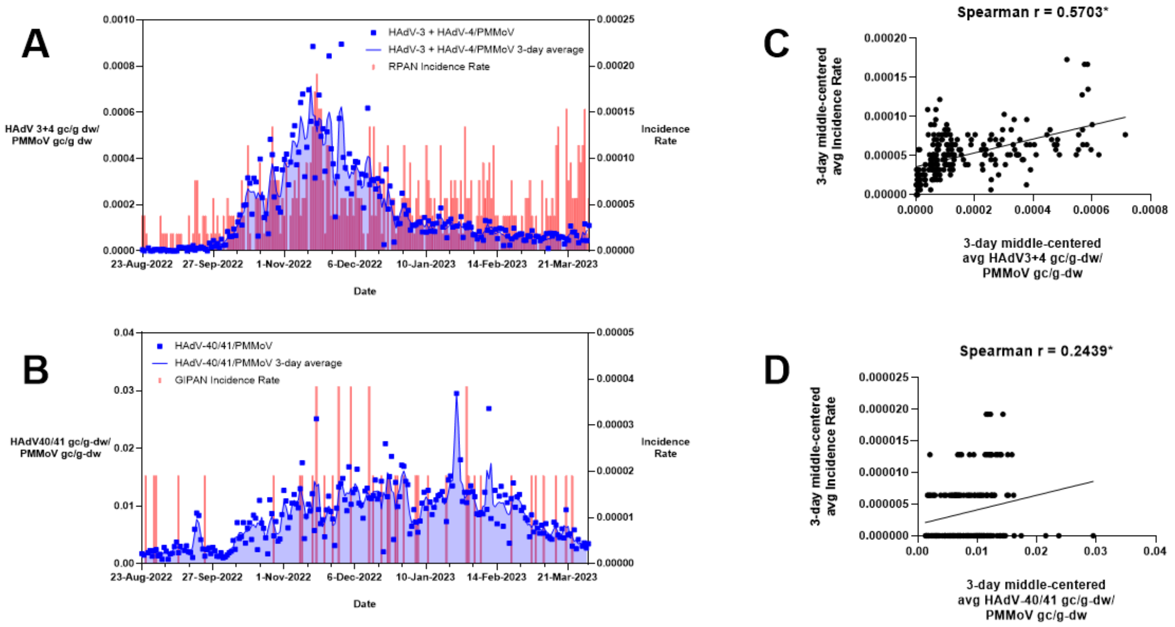


*Figure S6.* Comparison between wastewater and RPAN/GIPAN data. In panels A and B, blue squares represent human adenovirus targets in wastewater normalized by PMMoV and blue lines represent a 3-day middle-centered average with the area shaded under the curve. Wastewater data is plotted on the left y-axis. Red bar graphs represent clinical data shown as RPAN (respiratory panel) incidence rate and GIPAN (gastrointestinal panel) incidence rate for adenovirus positives based on a student population of 52,000. Clinical data is plotted on the right y-axis. All dates refer to specimen collection dates for wastewater and clinical specimens. In panels C and D, a corresponding correlation is shown for the data in panels A and B. The 3-day middle-centered average normalized human adenovirus targets in gene copies per gram dry weight were correlated with the 3-day middle-centered average incidence rate for the RPAN and GIPAN analyses. The Spearman's correlation coefficient was calculated and is noted on the correlation plot in panels C and D. Both correlation coefficients were significant.
